# Pediatric Venous Excess Ultrasound Score (P-VExUS): A Novel Approach to Assess Central Venous Pressure in the PICU

**DOI:** 10.64898/2026.02.11.26346088

**Authors:** Fernando de Lima Carioca, Nayara Hillebrand Franzon, Lívia da Silva Krzesinski, Isabel de Siqueira Ferraz, Roberto José Negrão Nogueira, Tiago Henrique de Souza

## Abstract

**Objectives:** To develop and validate pediatric adaptations of the Venous Excess Ultrasound Score (P-VExUS) for noninvasive estimation of central venous pressure (CVP) in critically ill children.

**Design:** Prospective observational study.

**Setting:** PICU of a tertiary-care teaching hospital.

**Patients:** Fifty-six mechanically ventilated children (median age 7.4 months, median weight 6.0 kg) with central venous catheters.

**Interventions:** None.

**Measurements and Main Results:** Venous Doppler ultrasonography of the inferior vena cava, hepatic, portal, and intrarenal veins was performed at the bedside. Two P-VExUS models were tested: (1) a categorical grading system (0–III) and (2) a semiquantitative point-based score (0–7). Both models showed significant associations with CVP. For predicting elevated CVP (>12 mmHg), model 1 achieved an AUROC of 0.74 (95% CI 0.61–0.85) with 45% sensitivity and 98% specificity, while model 2 demonstrated superior accuracy with an AUROC of 0.94 (95% CI 0.84–0.98), sensitivity 82%, and specificity 91% (*p* < 0.001). For detecting low CVP (<7 mmHg), model 2 also outperformed model 1 (AUROC 0.80 vs. 0.69, *p* = 0.02). Among individual venous Doppler components, intrarenal veins had the highest discriminative ability (AUROC 0.92), followed by hepatic (0.89) and portal (0.80) veins.

**Conclusions:** Two pediatric-specific P-VExUS models were feasible and accurate for estimating CVP in critically ill children. The point-based model (model 2) demonstrated superior diagnostic performance, supporting its potential as a noninvasive tool to assess venous congestion at the bedside.

**Research in Context:**

- Venous congestion, reflected by elevated central venous pressure (CVP), is associated with adverse outcomes in critically ill children, including mortality and renal dysfunction.
- The Venous Excess Ultrasound Score (VExUS) is validated in adults, but pediatric-specific adaptations and cutoff values remain poorly defined.
- There is a need for noninvasive, bedside tools to estimate CVP in children and guide fluid management in the PICU.

**What This Study Means:**

- This study validates pediatric-specific adaptations of the Venous Excess Ultrasound Score (P-VExUS) for estimating CVP in critically ill children.
- The semiquantitative point-based model provided more consistent and accurate discrimination of venous congestion compared with categorical grading.
- These findings highlight the feasibility and potential clinical utility of venous Doppler ultrasonography as a noninvasive bedside tool in the PICU.

## INTRODUCTION

Fluid tolerance is an emerging concept in the management of critically ill patients, including children, and refers to the ability to tolerate fluid administration without organ dysfunction. Central to the assessment of fluid tolerance and fluid overload syndrome is the monitoring of central venous pressure (CVP) (1, 2). It reflects the right atrial distension pressure relative to intrathoracic pressure and is influenced by factors such as ventricular afterload, cardiac tamponade, and positive-pressure ventilation (3). Although CVP is a poor marker of fluid responsiveness, it remains valuable for assessing venous congestion and fluid tolerance (4). In pediatrics, elevated CVP has been associated with several adverse outcomes, including increased mortality and renal dysfunction (5–9).

Accurate measurement of CVP can be challenging, as it relies on a patent, correctly positioned central venous catheter (CVC), which is itself prone to zeroing and calibration errors (10, 11). To overcome these technical limitations, a noninvasive approach to venous congestion assessment has been proposed based on ultrasonographic evaluation of multiple venous sites, including the inferior vena cava, hepatic vein, portal vein, and renal vein. This system, known as the Venous Excess Ultrasound (VExUS) grading system, demonstrated an association between venous congestion patterns and acute kidney injury in adult postoperative cardiac surgery patients (12). To date, however, few investigations have evaluated an analogous approach in the pediatric population (15–17).

Recently, we reported the diagnostic performance of hepatic vein Doppler for estimating CVP in mechanically ventilated children (18). Expanding this earlier work, the present study sought to develop pediatric-specific adaptations of the VExUS score (P-VExUS). Accordingly, the primary objective was to establish and characterize the P-VExUS score, while secondary objectives were to assess its diagnostic accuracy in identifying CVP values above 12 mmHg and below 7 mmHg — thresholds that have been associated with more favorable clinical outcomes (5, 8).

## METHODS

This prospective observational study was carried out between March 2023 and August 2025 in the Pediatric Intensive Care Unit (PICU) of the Clinical Hospital at the State University of Campinas (UNICAMP), São Paulo, Brazil. This study was performed in line with the principles of the Declaration of Helsinki. Approval was granted by the UNICAMP’s Research and Ethics Committee, approval # 67019422.0.0000.5404. Written informed consent was obtained from the parents.

### Participants

All invasively mechanically ventilated children aged 0 to 14 years with a CVC in place were screened for eligibility. Inclusion required appropriate CVC tip positioning—near the junction of the superior vena cava and right atrium—and a reliable CVP waveform. Exclusion criteria included: (1) interruption between the cava and right atrium; (2) inadequate acoustic window due to factors such as surgical wounds, dressings, or drainage tubes; (3) suspected or confirmed intra-abdominal hypertension; (4) structural abnormalities of the kidneys or liver; (5) hepatic cirrhosis or portal hypertension; (6) clinical evidence of respiratory distress; (7) presence of cardiac arrhythmias; and (8) unreliable CVP waveform.

### Study Protocol

Before ultrasound assessment, demographic and anthropometric data, primary diagnosis, vital signs, cardiac index, mean airway pressure, and mechanical ventilation parameters were recorded. After the examination, the CVP transducer was zeroed at the right atrial level, and the corresponding CVP value was documented.

### Ultrasound examinations

All ultrasound examinations were performed at the bedside by the same pediatric intensivist (THdS) using either a GE Vivid Q system with a 3.5–8 MHz or a Mindray DC-70 system with a 2.3–7.2 MHz phased-array transducer. Patients were examined in the supine position with the head of the bed elevated 15–30 degrees.

IVC diameters were measured in a subcostal sagittal view from the subxiphoid window, with maximum and minimum values obtained during a controlled respiratory cycle in B-mode, 1 cm distal to the hepatic vein confluence. From the same window, maximal abdominal aortic diameter was measured in the long axis. IVC variability indices were calculated as collapsibility (IVCCI = [IVCmax − IVCmin]/IVCmax), distensibility (IVCDI = [IVCmax − IVCmin]/IVCmin), and respiratory variability (ΔIVC = [IVCmax − IVCmin]/mean[IVCmax, IVCmin]). Ratios of IVCmax and IVCmin to aortic diameter (Ao) and body surface area (BSA) were also recorded (19).

Hepatic and portal vein Doppler examinations were performed according to previously described protocols (12, 20), with the transducer positioned longitudinally between the posterior and mid-axillary lines at the right ninth to tenth intercostal spaces (12, 20). Hepatic vein Doppler (HVD) was classified as normal (S ≥ D), mildly abnormal (S < D), or severely abnormal (reversed S wave). Portal vein Doppler was used to calculate the portal pulsatility index (PPI), defined as PPI (%) = 100 × [(Vmax − Vmin) / Vmax], based on peak and nadir velocities over the cardiac cycle.

Given site-dependent variability in venous Doppler signals, venous congestion was classified as follows: normal pattern—continuous flow or an S wave equal to or greater than the D wave; mildly abnormal—D wave greater than S wave; and severely abnormal—D wave exceeding S wave by more than 50%.

To minimize respirophasic effects on venous waveforms, Doppler acquisitions were obtained during expiration, repeated three times, and averaged for analysis. The ultrasound operator was blinded to the CVP measurement at the time of Doppler acquisition, with CVP values recorded only after completion of the ultrasound examination. Blinding to other clinical parameters was not feasible, as these were readily available at the bedside as part of standard clinical care.

### Statistical Analysis

Statistical analyses were performed using MedCalc Statistical Software version 14.8.1 (MedCalc Software bvba, Ostend, Belgium). Data distribution was assessed with the Kolmogorov–Smirnov and Shapiro–Wilk tests. Continuous nonparametric variables are reported as median (IQR), and categorical variables as counts and percentages.

As no pediatric-specific VExUS score has yet been validated, we sought to determine optimal cutoff values for IVC–derived parameters and the PPI to identify CVP >12 mmHg. Receiver operating characteristic (ROC) curve analysis was performed for this purpose, and optimal thresholds were identified using Youden’s J statistic in univariate analyses.

Two complementary ROC analyses were performed: one evaluating the discriminative performance of continuous Doppler-derived variables (IVC-derived measurements and PPI) and identifying optimal cutoff values, and another assessing diagnostic performance after transforming these variables into ordinal score components according to the predefined P-VExUS grading framework. Previously proposed age-specific reference thresholds for IVC-derived indices were also tested to evaluate their performance in our cohort (21, 22). Based on these analyses, two adapted P-VExUS models were evaluated for their ability to predict elevated CVP:

- **Model 1 (Graded Classification):** Adapted from the original grading system proposed by Beaubien-Souligny et al., patients were classified into P-VExUS grades 0 to III. Grade 0 was assigned when the IVC measurement was below the identified cutoff value. If the IVC exceeded this cutoff, patients were categorized according to venous Doppler abnormalities: Grade I for normal or mildly abnormal findings in any venous territory; Grade II for a severely abnormal Doppler pattern in a single venous site; and Grade III for severely abnormal patterns in at least two venous territories.
- **Model 2 (Point-Based Score):** Each site was assigned a score: 0 for normal findings, 1 point for an abnormal IVC, and 1 or 2 points for mildly or severely abnormal Doppler patterns, respectively. The total score ranged from 0 to 7.

The accuracy of both models for predicting CVP >12 mmHg and <7 mmHg was assessed using ROC analysis, with AUCs compared by DeLong’s method for correlated curves (23).

Based on prior data, we expected that 20% of the study population would have a CVP >12 mmHg (8). To identify a 0.30 difference between the AUROC of the P-VExUS score and the null hypothesis value of 0.50 (indicating no discriminatory ability), we calculated that a sample of 55 measurements would be required, with a power of 90% and a type I error rate of 5%.

## RESULTS

During the study period, 66 patients were eligible for inclusion. Four patients were excluded due to insufficient acoustic windows caused by dressings in the thoraco-abdominal region, while six patients were excluded due to unreliable CVP measurements. Therefore, a total of 56 patients were enrolled. Among these, 38 had participated in the earlier hepatic Doppler study; however, only those with complete multi-territory venous Doppler datasets were included in the present analysis (18). Most patients were admitted for acute respiratory failure (38%), post-cardiac surgery care (28%), or sepsis (25%). Among postoperative congenital heart disease patients, none had univentricular physiology or severe tricuspid regurgitation. Demographic characteristics of the study population can be found in ***Table 1***.

**Table 1.**
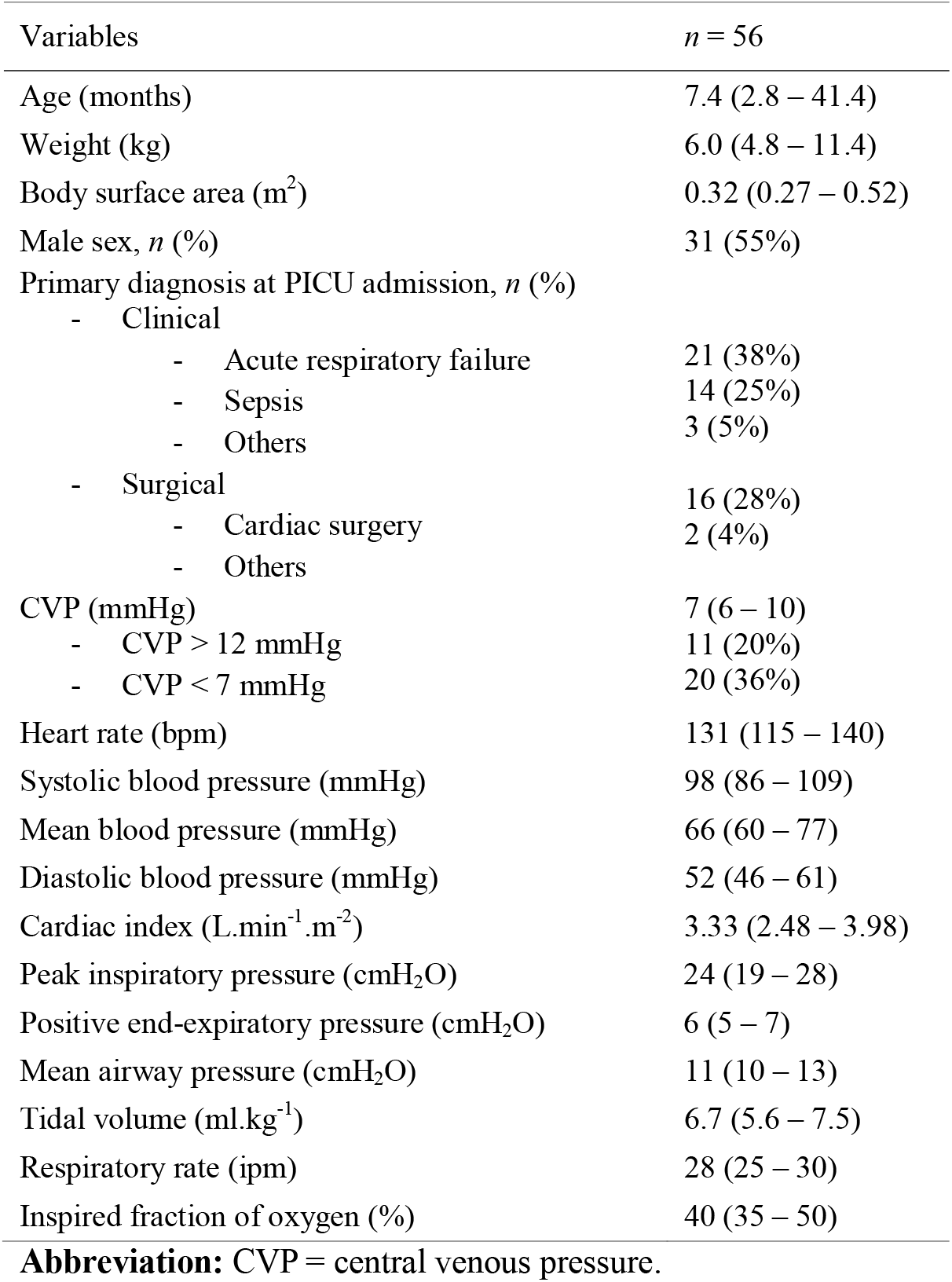
Demographic and clinical characteristics of pediatric patients included in the study, expressed as median (interquartile range) for continuous variables and absolute numbers (percentages) for categorical variables.

Among all IVC-derived indices, only the IVCmax/BSA ratio was able to identify a CVP > 12 mmHg with a statistically significant area under the ROC curve (AUROC 0.68 [95% CI 0.54–0.80], *p* = 0.041) (Table S1, Online Supplement). The optimal cutoff value was 1.96 cm/m^2^. Therefore, in P-VExUS model 1, patients with IVCmax/BSA < 1.96 cm/m^2^ were classified as having a normal pattern, while in model 2, patients with IVCmax/BSA > 1.96 cm/m^2^ received 1 point.

For PVD, the PPI showed good accuracy for detecting CVP >12 mmHg (AUROC 0.89, 95% CI 0.78–0.96, *p* < 0.001). ROC analysis identified 35% as the optimal threshold; however, this differed minimally from the conventional 30% cutoff, likely reflecting inherent variability in Doppler-derived portal flow. When PVD scores were defined using either threshold (with ≥50% consistently assigned 2 points), ROC performance was similar (AUROC 0.81 vs. 0.80), with no significant difference between curves (*p* > 0.05).

To preserve consistency with established VExUS grading and facilitate comparability across studies, we therefore retained the traditional 30% and 50% pulsatility thresholds. Accordingly, PVD was classified as normal when minimal variation in flow velocity was present during the cardiac cycle (PPI < 30%), mildly abnormal when PPI was 30–50%, and severely abnormal when PPI was ≥ 50%. Representative examples of each pattern are shown in ***Figure 1***, and the classification frameworks for P-VExUS models 1 and 2 are summarized in ***Table 2***.

**Table 2.**
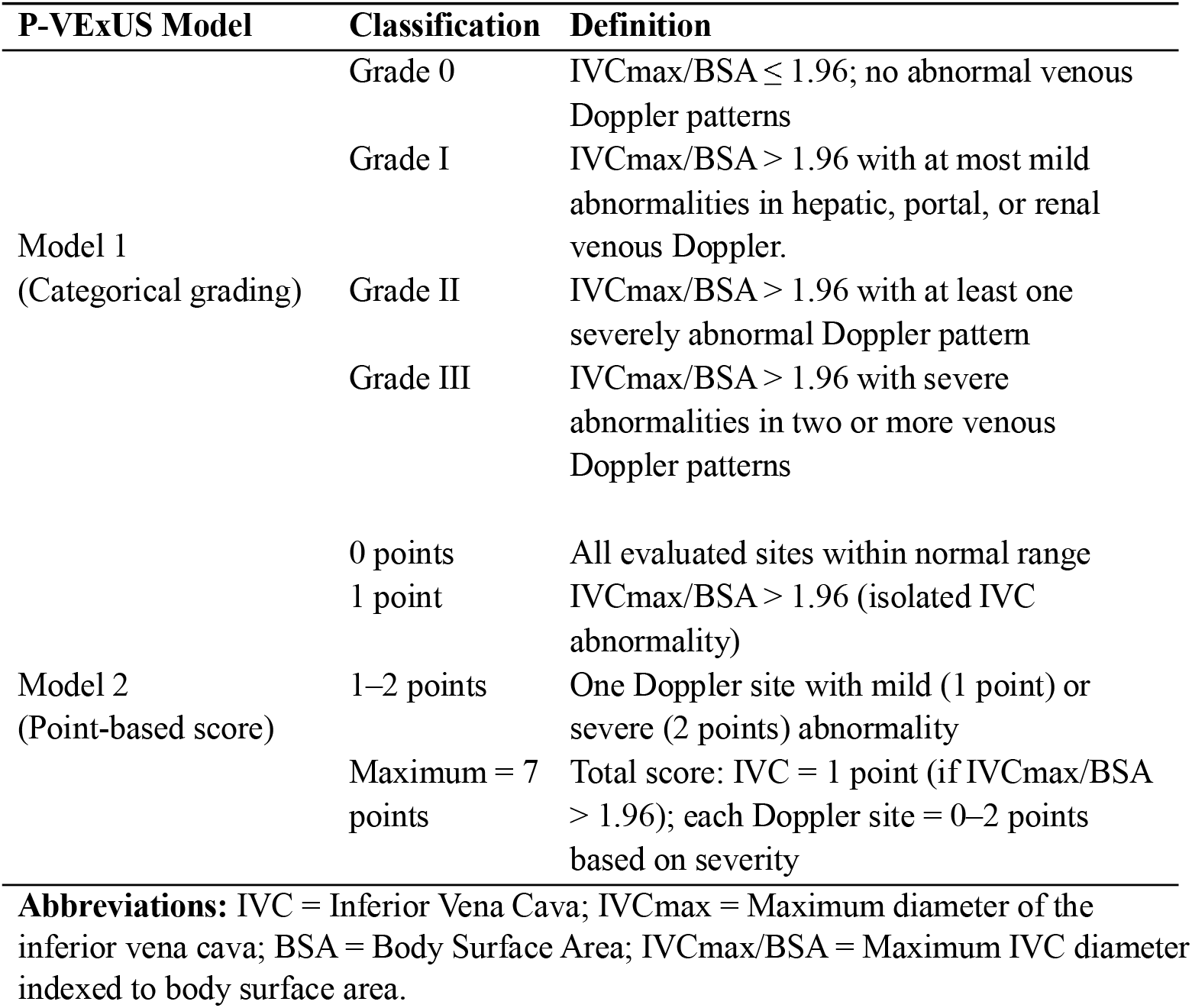
Classification Criteria for P-VExUS Models 1 and 2.

**Figure 1.**
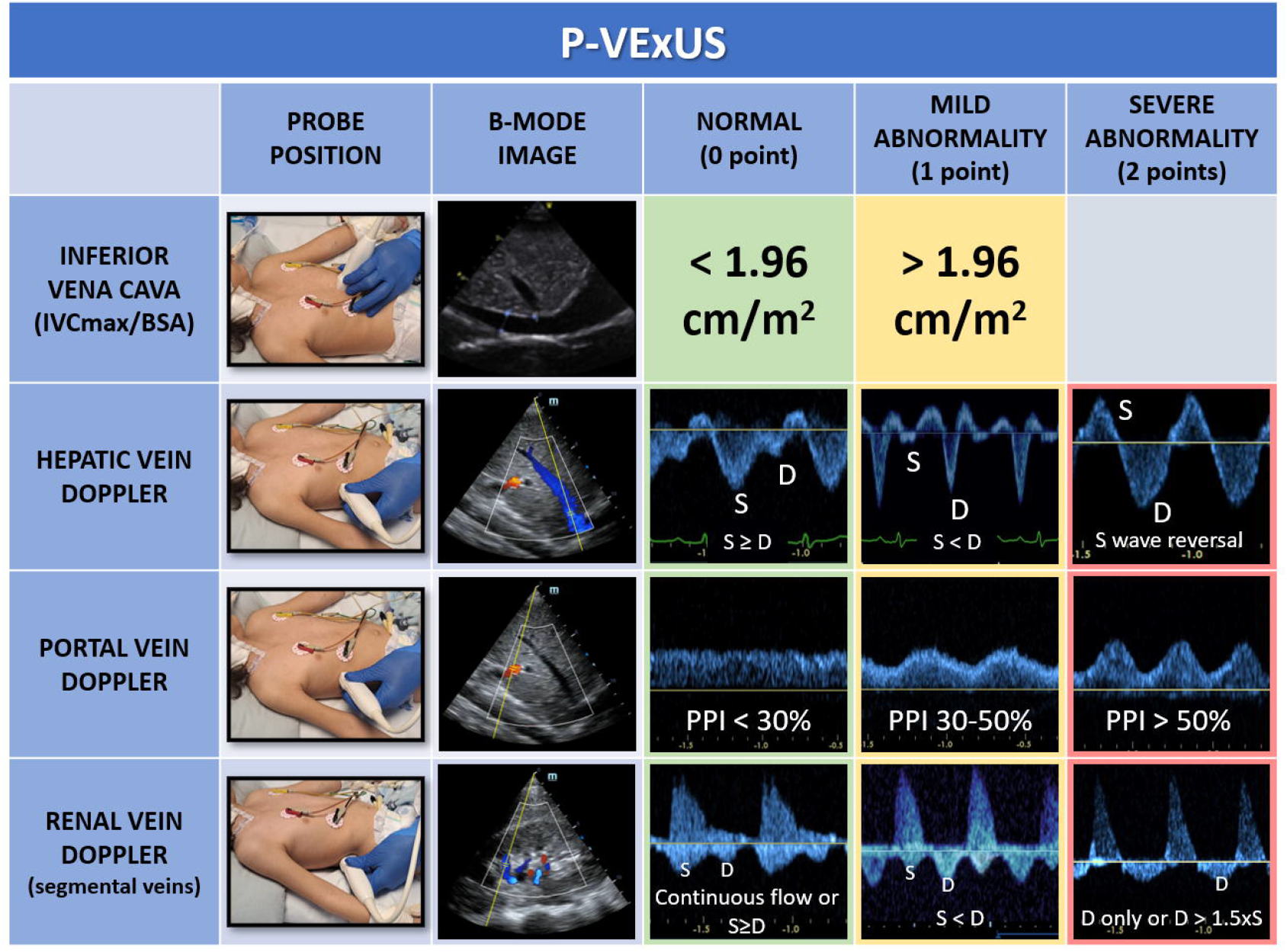
Schematic representation of the P-VExUS framework illustrating, for each component, the transducer position, the corresponding B-mode image, and the Doppler waveform patterns classified as normal (0 point), mild abnormality (1 point), or severe abnormality (2 points). Inferior vena cava assessment is based on the body surface area– indexed maximal diameter (IVCmax/BSA), with values <1.96 cm/m^2^ considered normal and values >1.96 cm/m^2^ indicating abnormal dilation. Hepatic vein Doppler patterns progress from a normal systolic-dominant waveform (S ≥ D), to a mildly abnormal pattern with diastolic predominance (S < D), and to severe abnormality characterized by systolic flow reversal. Portal vein Doppler is graded according to the portal pulsatility index (PPI), ranging from minimal pulsatility (PPI < 30%) to moderate pulsatility (PPI 30–50%) and severe pulsatility (PPI > 50%). Renal vein Doppler (segmental veins) evolves from continuous flow or systolic-dominant patterns (S ≥ D), to discontinuous flow with diastolic predominance (S < D), and finally to severe congestion marked by monophasic diastolic flow or marked diastolic dominance (D > 1.5 × S). The integrated P-VExUS score reflects the severity of systemic venous congestion in critically ill pediatric patients.

CVP increased progressively with higher congestion categories in both P-VExUS models, with a more linear and consistent relationship observed in model 2 (***Figure 2***).

**Figure 2.**
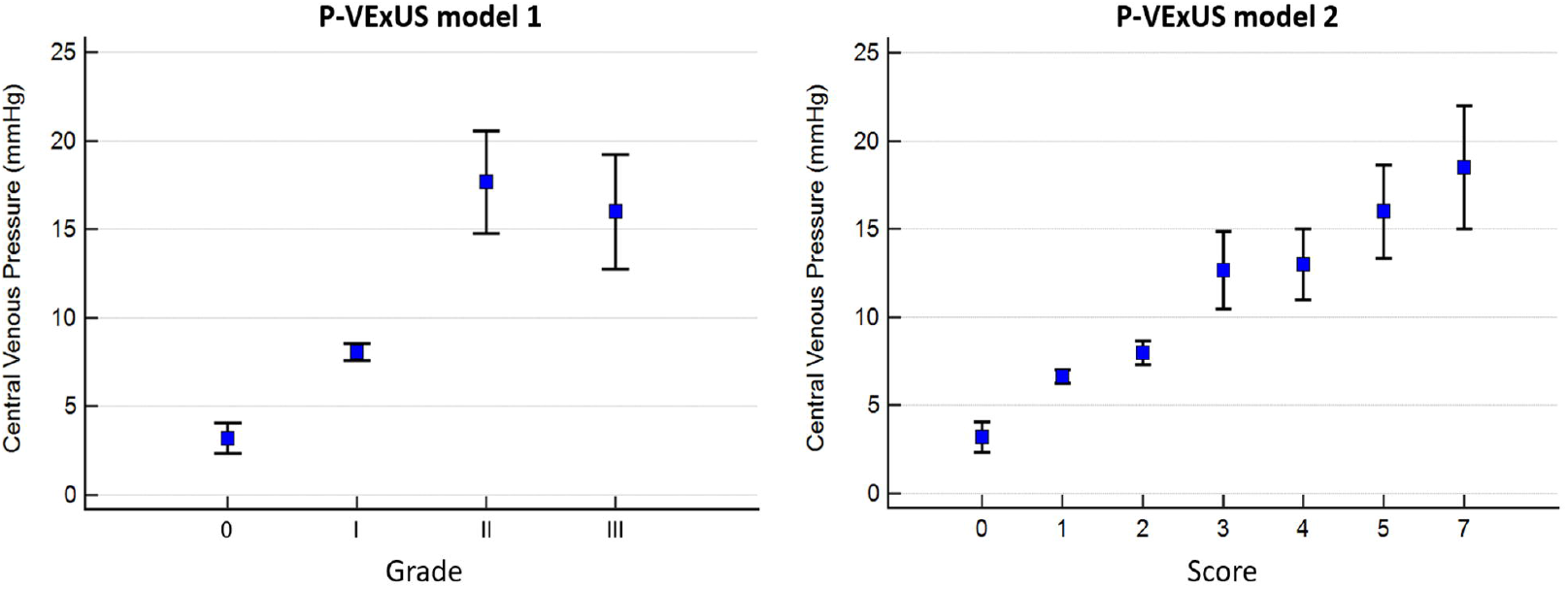
Central venous pressure (CVP) values according to P-VExUS model 1 (left) and P-VExUS model 2 (right). CVP is expressed as mean ± standard error of the mean (SEM) for each congestion grade (model 1) or score (model 2). Both models show a progressive increase in CVP with higher congestion categories.

Both P-VExUS models demonstrated diagnostic accuracy for identifying CVP >12 mmHg, with model 2 showing superior performance (*p* < 0.001). In P-VExUS model 1, the presence of Grade II or higher congestion yielded an AUROC of 0.74 (95% CI 0.61–0.85, *p* < 0.001), with a sensitivity of 45% and a specificity of 98%. The P-VExUS model 2, using a threshold of ≥ 3 points, demonstrated excellent discrimination, with an AUROC of 0.94 (95% CI 0.84–0.98, *p* < 0.001). This model achieved a sensitivity of 82% and specificity of 91% (***Table 3***).

**Table 3.** Diagnostic accuracy of P-VExUS models for predicting central venous pressure extremes (>12 mmHg and <7 mmHg), with values in parentheses indicating 95% confidence intervals.

| Parameter | P-VExUS Model 1<br>(Categorical grading) | P-VExUS Model 2<br>(Point-based score) |
| --- | --- | --- |
| **Prediction of CVP >12 mmHg** |  |  |
| AUROC (95% CI) | 0.74 (0.61–0.85) | 0.94 (0.84–0.98) |
| Optimal cutoff | ≥ Grade II | ≥3 points |
| Sensitivity (%) | 45 (17 – 77) | 82 (48 – 98) |
| Specificity (%) | 98 (88 – 100) | 91 (79 – 97) |
| **Prediction of CVP <7 mmHg** |  |  |
| AUROC (95% CI) | 0.69 (0.55–0.81) | 0.80 (0.67–0.89) |
| Optimal cutoff | Grade 0 | ≤1 point |
| Sensitivity (%) | 25 (9 – 49) | 75 (51 – 91) |
| Specificity (%) | 100 (90 – 100) | 67 (49 – 81) |

**Abbreviations:** CVP = central venous pressure; AUROC = area under the receiver operating characteristic curve.

When evaluating the performance of individual venous Doppler score components for detecting elevated CVP (>12 mmHg), the renal vein score showed the highest discriminative ability, with an AUROC of 0.92 (95% CI 0.82–0.98), followed by the hepatic vein score (AUROC 0.89, 95% CI 0.77–0.96) and the portal vein score (AUROC 0.80, 95% CI 0.67–0.89). The IVC score demonstrated the lowest discriminative performance, with an AUROC of 0.56 (95% CI 0.42–0.69). There was no statistically significant difference between the AUROC of P-VExUS model 2 and the renal vein score (*p* = 0.63).

For the identification of low CVP (< 7 mmHg), P-VExUS model 2 showed better diagnostic performance compared to model 1 (*p* = 0.02). P-VExUS model 2 had an AUROC of 0.80 (95% CI 0.67–0.89, *p* < 0.001), with a cutoff value ≤ 1, yielding a sensitivity of 75% and a specificity of 67%. In contrast, P-VExUS model 1 demonstrated an AUROC of 0.69 (95% CI 0.55–0.81, *p* < 0.001), using Grade 0 as the cutoff, with a sensitivity of 25% and specificity of 100%. Detailed results regarding the diagnostic performance of both P-VExUS models and each venous Doppler site are available in the **Online Supplement**.

## DISCUSSION

This prospective study proposes two novel pediatric adaptations of the Venous Excess Ultrasound Score (P-VExUS) to noninvasively estimate CVP in critically ill children. Our findings demonstrate that both models, particularly the point-based P-VExUS model 2, were able to discriminate between low and high CVP states with moderate to high accuracy. Notably, model 2 showed superior performance for both extremes of CVP, with AUROCs of 0.80 and 0.93 for predicting CVP <7 mmHg and >12 mmHg, respectively.

Interest in VExUS has expanded notably as emerging evidence underscores the limitations of IVC ultrasound in evaluating systemic venous congestion. Despite widespread use, IVC-derived indices have performed poorly in predicting CVP or fluid responsiveness in both adults and critically ill children (24–26). These limitations are accentuated in pediatrics due to smaller anatomic dimensions, developmental variability, and respiratory displacement (27). A recent multicenter study in mechanically ventilated children confirmed that IVC diameters, collapsibility and distensibility indices, and the IVC-to-aorta ratio did not correlate with invasively measured CVP (25). Notably, the limitations of IVC ultrasonography appear to be especially prominent in mechanically ventilated pediatric patients (28).

Our findings are consistent with this body of evidence. We observed that none of the traditional IVC-derived parameters reliably identified elevated CVP, with the single exception of IVCmax/BSA. Because IVC size is highly dependent on somatic growth, indexing to BSA may offer conceptual advantages. Adult data further support this concept. In Taniguchi et al.’s cohort, the optimal threshold for maximal IVC diameter to identify right atrial pressure ≥10 mmHg increased from 17 mm in patients with BSA <1.61 m^2^ to 21 mm in those with BSA ≥1.61 m^2^, highlighting the substantial physiologic impact of body size on IVC-derived estimates of venous pressure (29). However, the pediatric evidence base necessary to support the routine use of size-indexed IVC thresholds remains notably sparse and inconsistent (19).

Although pediatric data remain limited, early studies suggest a potential role for multisite venous Doppler ultrasonography in assessing systemic congestion in children. In the pioneering study by Menéndez-Suso et al., one of the first to systematically apply VExUS in a pediatric population, the score showed a strong correlation with CVP in 33 children, with median CVP values demonstrating a clear stepwise increase across congestion grades (30). Natraj et al. further expanded the clinical relevance of VExUS by showing that a score >2 was strongly associated with acute kidney injury in children with postoperative right ventricular dysfunction (sensitivity 83%, specificity 84%); notably, improvements in VExUS paralleled reductions in CVP and fluid balance following targeted therapy (16).

More recently, a post hoc analysis by Siuba et al. showed a moderately strong negative association between VExUS and right ventricular–pulmonary arterial (RV–PA) coupling, with higher VExUS grades aligning with progressively impaired right ventricular longitudinal shortening relative to pulmonary arterial systolic pressure (17). These findings reinforce that abnormalities in abdominal venous Doppler waveforms reflect not only venous congestion but also the hemodynamic consequences of right-sided cardiac dysfunction.

While these foundational studies highlight the promise of VExUS in pediatrics, they did not address developmental or physiologic distinctions unique to this population. Aside from applying IVC thresholds derived from small pediatric cohorts, most components of the score were directly extrapolated from adult frameworks. A key strength of our study is the data-driven calibration of venous ultrasonography to invasively measured CVP, rather than reliance on fixed thresholds adapted from adult practice. Using ROC analyses and Youden’s index, we derived two optimized cutoffs— IVCmax/BSA >1.96 cm/m^2^ and PPI ≥35%—and integrated these values into graded and point-based pediatric VExUS models. Notably, a recent large pediatric cohort following congenital heart surgery identified a similar optimal portal pulsatility threshold (PPI >30.3%) associated with adverse outcomes, lending external support to the range identified in our analysis (31). After confirming that PPI ≥30% demonstrated diagnostic performance comparable to the ≥ 35% threshold, we elected to retain the 30% cutoff to preserve continuity with established adult criteria and facilitate clinical adoption in pediatrics.

Interestingly, our proposed P-VExUS model 2 incorporates a graded, semiquantitative framework that allows for greater granularity compared to the categorical grading of model 1. This more refined scoring system produced a progressive and linear increase in CVP values across score categories and demonstrated better performance in identifying both high and low CVP thresholds. These results suggest that pediatric-specific adaptations of the VExUS score may enhance its clinical applicability and diagnostic value in younger populations.

Our findings regarding the diagnostic performance of individual venous Doppler components also raise important considerations about the potential simplification of VExUS in pediatric practice. Notably, renal vein Doppler demonstrated the highest discriminative ability in our cohort (AUROC 0.91). Similar observations have been reported by other investigators (16, 30). Together, these findings suggest that renal venous Doppler may carry substantial diagnostic weight and raise the possibility that simplified approaches focused on key venous territories could retain meaningful clinical utility in children. Other venous sites also warrant investigation in pediatric populations, including the femoral veins, which have shown promising results in adult studies (32, 33).

However, important practical considerations must be acknowledged. Assessing intrarenal veins is often technically challenging, particularly in smaller children, and Doppler waveforms may vary according to the specific location within the renal vasculature (34). These sources of variability highlight the need for standardized acquisition protocols and clearer methodological reporting. To optimize feasibility and bedside reproducibility, our study focused on Doppler interrogation of the segmental veins of the right kidney. Whether bilateral renal venous assessment provides incremental diagnostic value remains unknown and warrants further investigation. Although renal venous Doppler shows promise as a simplified surrogate of systemic venous congestion, its clinical applicability will require careful prospective validation.

Our study has limitations. First, the sample size was relatively small, and data were collected at a single center, with all ultrasound examinations performed by a single experienced operator. Future studies should include structured evaluations of inter- and intraobserver reproducibility among clinicians with varying levels of POCUS training, as well as assessments of the time required to complete the ultrasound examination. Second, only invasively mechanically ventilated children were included, and our findings should not be extrapolated to spontaneously breathing patients. Additionally, although CVP was used as the reference standard, it remains an imperfect surrogate for true intravascular volume status or organ-level congestion. Larger, multicenter studies are needed to determine the prognostic value of P-VExUS and to assess whether its use can guide therapeutic decisions.

## CONCLUSIONS

In this prospective study, we evaluated two pediatric adaptations of the VExUS grading system for noninvasive assessment of CVP in critically ill children. Both models correlated with invasively measured CVP, supporting P-VExUS—particularly model 2—as a structured bedside approach to venous congestion assessment in the PICU. Larger multicenter studies are needed to validate these findings and define the clinical role of P-VExUS in pediatric fluid management.

## Acknowledgments

We thank the attending physicians, resident physicians, nursing staff, and legal guardians of the participants in this study.

## Author Contributions

Conception and design: Tiago H. de Souza; Acquisition of data: Fernando de Lima Carioca, Nayara Hillebrand Franzon, Lívia da Silva Krzesinski, and Tiago H. de Souza; Analysis and interpretation of data: Fernando de Lima Carioca and Tiago H. de Souza; Drafting the article: Fernando de Lima Carioca and Tiago H. de Souza; Revising the article critically for important intellectual content: Isabel de S. Ferraz, and Roberto J. N. Nogueira; All of the authors read and approved the manuscript.

## Funding

The authors declare that no funds, grants, or other support were received during the preparation of this manuscript.

## Competing Interests

The authors have no competing interests to declare that are relevant to the content of this article.

## Data availability statement

The data that support the findings of this study are available from the corresponding author upon reasonable request.

## Consent statement

This study was performed in line with the principles of the Declaration of Helsinki. Approval was granted by the UNICAMP’s Research and Ethics Committee, approval # 67019422.0.0000.5404. Written informed consent was obtained from the parents.

## Notes

### Competing Interest Statement

The authors have declared no competing interest.

