## Supplementary material for "Pediatric Venous Excess Ultrasound Score (P-VExUS): A Novel Approach to Assess Central Venous Pressure in the PICU": Online Supplement

**Summary**

[**Table S4.** Diagnostic accuracy of P-VExUS models for predicting low central venous pressure (<7 mmHg). Values in parentheses indicate 95% confidence intervals.	3](#_Toc217293059)

[**Table S5.** Diagnostic accuracy of individual venous Doppler sites for predicting low central venous pressure (<7 mmHg). Values in parentheses indicate 95% confidence intervals.	3](#_Toc217293060)

[**Figure S3.** Receiver operating characteristic (ROC) curves for predicting low central venous pressure (CVP < 7 mmHg) using two pediatric Venous Excess Ultrasound Score (P-VExUS) models	6](#_Toc217293063)

[**Figure S4.** Receiver operating characteristic (ROC) curves of individual venous Doppler components for predicting low central venous pressure (CVP < 7 mmHg)	7](#_Toc217293064)

### **Table S1.** Diagnostic performance (AUROC) of inferior vena cava–derived parameters for predicting central venous pressure >12 mmHg.

| **IVC-derived parameter** | **AUROC** | **95% CI** | **p-value** |
| --- | --- | --- | --- |
| IVC max/Ao | 0,520 | 0,383 to 0,656 | 0,817 |
| IVC min/Ao | 0,535 | 0,397 to 0,670 | 0,704 |
| IVC max/BSA | **0,682** | **0,544 to 0,800** | **0,041** |
| IVC min/BSA | 0,537 | 0,399 to 0,672 | 0,718 |
| IVCDI | 0,532 | 0,394 to 0,667 | 0,750 |
| IVCCI | 0,532 | 0,394 to 0,667 | 0,750 |
| ΔVCI | 0,532 | 0,394 to 0,667 | 0,750 |
| Mannarino et al.* | 0,586 | 0,446 to 0,716 | 0,282 |
| Kathuria et al.** | 0,518 | 0,381 to 0,654 | 0,829 |

***Abbreviations:*** *IVC max = maximum inferior vena cava diameter; IVC min = minimum inferior vena cava diameter; Ao = aorta; BSA = body surface area; IVCDI = IVC distensibility index; IVCCI = IVC collapsibility index; ΔIVC = respiratory variation in IVC diameter; AUROC = area under the receiver operating characteristic curve; 95% CI = 95% confidence interval; CVP = central venous pressure.*

***Age-dependent reference values reported in:***

** Mannarino S, Bulzomì P, Codazzi AC, et al. Inferior vena cava, abdominal aorta, and IVC-to-aorta ratio in healthy Caucasian children: Ultrasound Z-scores according to BSA and age. J Cardiol. 2019;74:388–393.
** Kathuria N, Ng L, Saul T, et al. The baseline diameter of the inferior vena cava measured by sonography increases with age in normovolemic children. J Ultrasound Med. 2015;34:1091–1096.*

### **Table S2.** Diagnostic accuracy of P-VExUS models for predicting elevated central venous pressure (>12 mmHg). Values in parentheses indicate 95% confidence intervals.

| **Parameter** | **P-VExUS Model 1** | **P-VExUS Model 2** |
| --- | --- | --- |
| AUROC | 0.74 (0.61 – 0.85) | 0.94 (0.84 – 0.98) |
| Cut-off | ≥ Grade II | ≥3 |
| Sensitivity | 45 (17 – 77) | 82 (48 – 98) |
| Specificity | 98 (88 – 100) | 91 (79 – 97) |
| +LR | 20.4 (2.7 – 157.8) | 9.2 (3.5 – 24.4) |
| -LR | 0.56 (0.3 – 1.0) | 0.2 (0.06 – 0.7) |
| +PV (%) | 83 (39.3 – 97.5) | 69 (46 – 86) |
| -PV (%) | 88 (81 – 92) | 95 (85 – 99) |

***Abbreviations:*** *P-VExUS = Pediatric Venous Excess Ultrasound Score; CVP = central venous pressure; AUROC = area under the receiver operating characteristic curve; +LR = positive likelihood ratio; -LR = negative likelihood ratio; +PV = positive predictive value; -PV = negative predictive value.*

### **Table S3.** Diagnostic accuracy of individual venous Doppler sites for predicting elevated central venous pressure (>12 mmHg). Values in parentheses indicate 95% confidence intervals.

| **Parameter** | **IVC** | **Hepatic Vein** | **Portal Vein** | **Renal Vein** |
| --- | --- | --- | --- | --- |
| AUROC | 0.55 (0.42 – 0.69) | 0.89 (0.77 – 0.96) | 0.80 (0.67 – 0.89) | 0.92 (0.82 – 0.98) |
| Cut-off | >0 | >0 | >0 | >0 |
| Sensitivity | 100 (71 – 100) | 91 (56 – 100) | 82 (48 – 98) | 91 (59 – 100) |
| Specificity | 11 (4 – 24) | 84 (70 – 93) | 69 (53 – 82) | 93 (82 – 99) |
| +LR | 1.12 (1.0 – 1.2) | 5.84 (2.9 – 11.8) | 2.63 (1.6 – 4.4) | 13.6 (4.5 – 41.3) |
| -LR | 0 | 0.11 (0.02 – 0.7) | 0.26 (0.07 – 0.9) | 0.097 (0.02 – 0.6) |
| +PV (%) | 22 (20 – 23.4) | 59 (41 – 74) | 39 (28 – 52) | 77 (52 – 91) |
| -PV (%) | 100 | 97 (85 – 100) | 94 (81 – 98) | 98 (87 – 100) |

***Abbreviations:*** *CVP = central venous pressure; AUROC = area under the receiver operating characteristic curve; +LR = positive likelihood ratio; -LR = negative likelihood ratio; +PV = positive predictive value; -PV = negative predictive value.*

### **Table S4.** Diagnostic accuracy of P-VExUS models for predicting low central venous pressure (<7 mmHg). Values in parentheses indicate 95% confidence intervals.

| **Parameter** | **P-VExUS Model 1** | **P-VExUS Model 2** |
| --- | --- | --- |
| AUROC | 0.69 (0.55–0.81) | 0.80 (0.67–0.89) |
| Cut-off | Grade 0 | ≤1 point |
| Sensitivity | 25 (9 – 49) | 75 (51 – 91) |
| Specificity | 100 (90 – 100) | 67 (49 – 81) |
| +LR | - | 2.25 (1.33 – 3.81) |
| -LR | 0.75 (0.58 – 0.97) | 0.38 (0.17 – 0.83) |
| +PV (%) | 100 | 56 (42 – 68) |
| -PV (%) | 71 (65 – 76) | 83 (68 – 91) |

***Abbreviations:*** *P-VExUS = Pediatric Venous Excess Ultrasound Score; CVP = central venous pressure; AUROC = area under the receiver operating characteristic curve; +LR = positive likelihood ratio; -LR = negative likelihood ratio; +PV = positive predictive value; -PV = negative predictive value.*

### **Table S5.** Diagnostic accuracy of individual venous Doppler sites for predicting low central venous pressure (<7 mmHg). Values in parentheses indicate 95% confidence intervals.

| **Parameter** | **IVC** | **Hepatic Vein** | **Portal Vein** | **Renal Vein** |
| --- | --- | --- | --- | --- |
| AUROC | 0.62 (0.48 – 0.75) | 0.70 (0.56 – 0.81) | 0.64 (0.50 – 0.76) | 0.68 (0.54 – 0.80) |
| Cut-off | ≤0 | ≤0 | ≤0 |  |
| Sensitivity | 25 (9 – 49) | 95 (75 – 100) | 75 (51 – 91) | 100 (83 – 100) |
| Specificity | 100 (90 – 100) | 44 (28 – 62) | 50 (33 – 67) | 36 (21 – 54) |
| +LR | - | 1.71 (1.26 – 2.33) | 1.50 (0.99 – 2.27) | 1.57 (1.22 – 2) |
| -LR | 0.75 (0.58 – 0.97) | 0.11 (0.02 – 0.79) | 0.50 (0.22 – 1.14) | - |
| +PV (%) | 100 | 49 (41 – 56) | 45 (35 – 56) | 46 (40 – 53) |
| -PV (%) | 71 (65 – 76) | 94 (70 – 99) | 78 (61 – 89) | 100 |

***Abbreviations:*** *CVP = central venous pressure; AUROC = area under the receiver operating characteristic curve; +LR = positive likelihood ratio; -LR = negative likelihood ratio; +PV = positive predictive value; -PV = negative predictive value.*

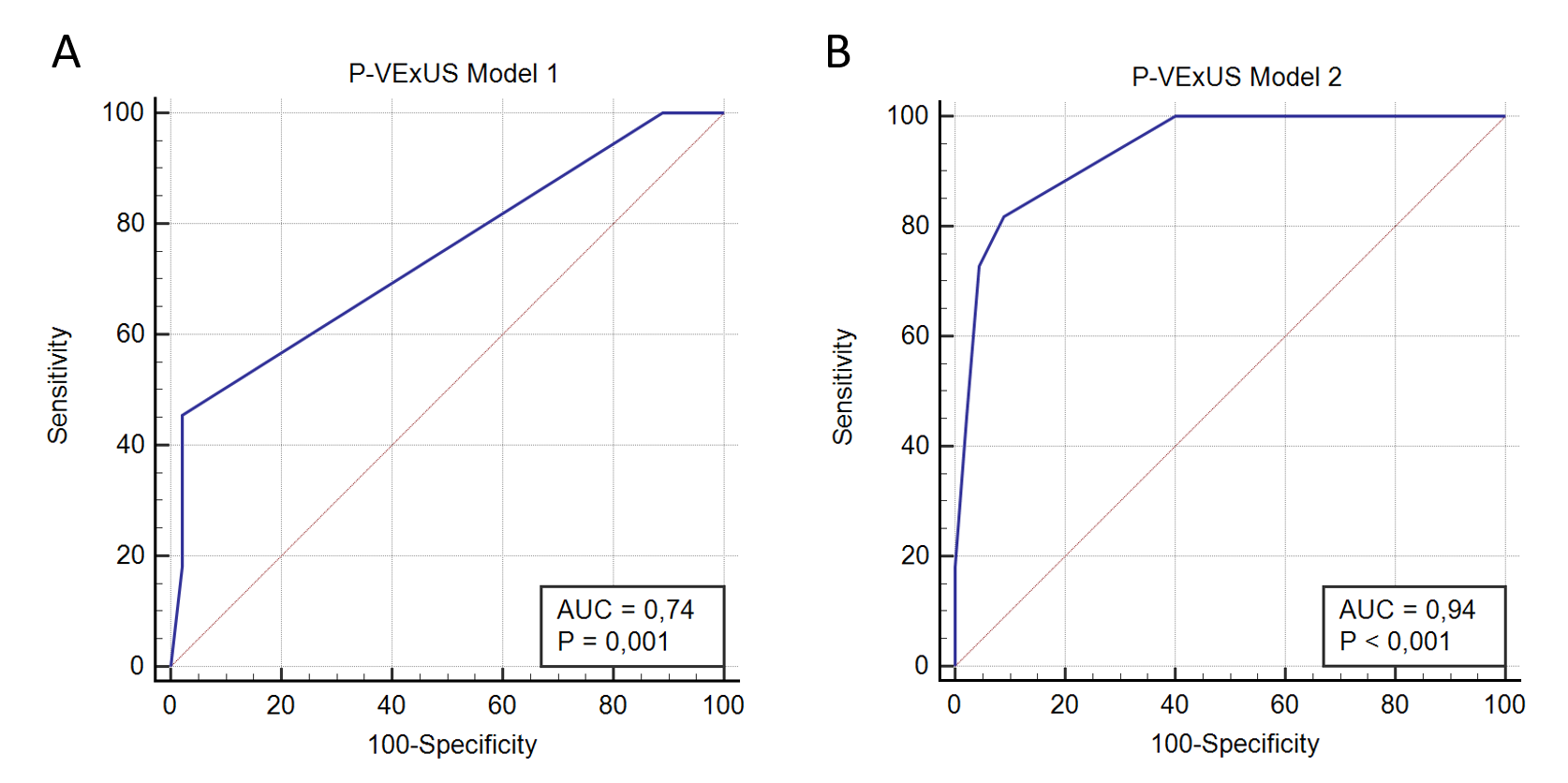

### **Figure S1. Receiver operating characteristic (ROC) curves of P-VExUS models for predicting elevated central venous pressure (CVP > 12 mmHg).** (A) P-VExUS Model 1 (categorical grading) demonstrated an AUROC of 0.74 (95% CI 0.61–0.85, p = 0.001). (B) P-VExUS Model 2 (point-based score) showed superior diagnostic accuracy, with an AUROC of 0.94 (95% CI 0.84–0.98, p < 0.001).

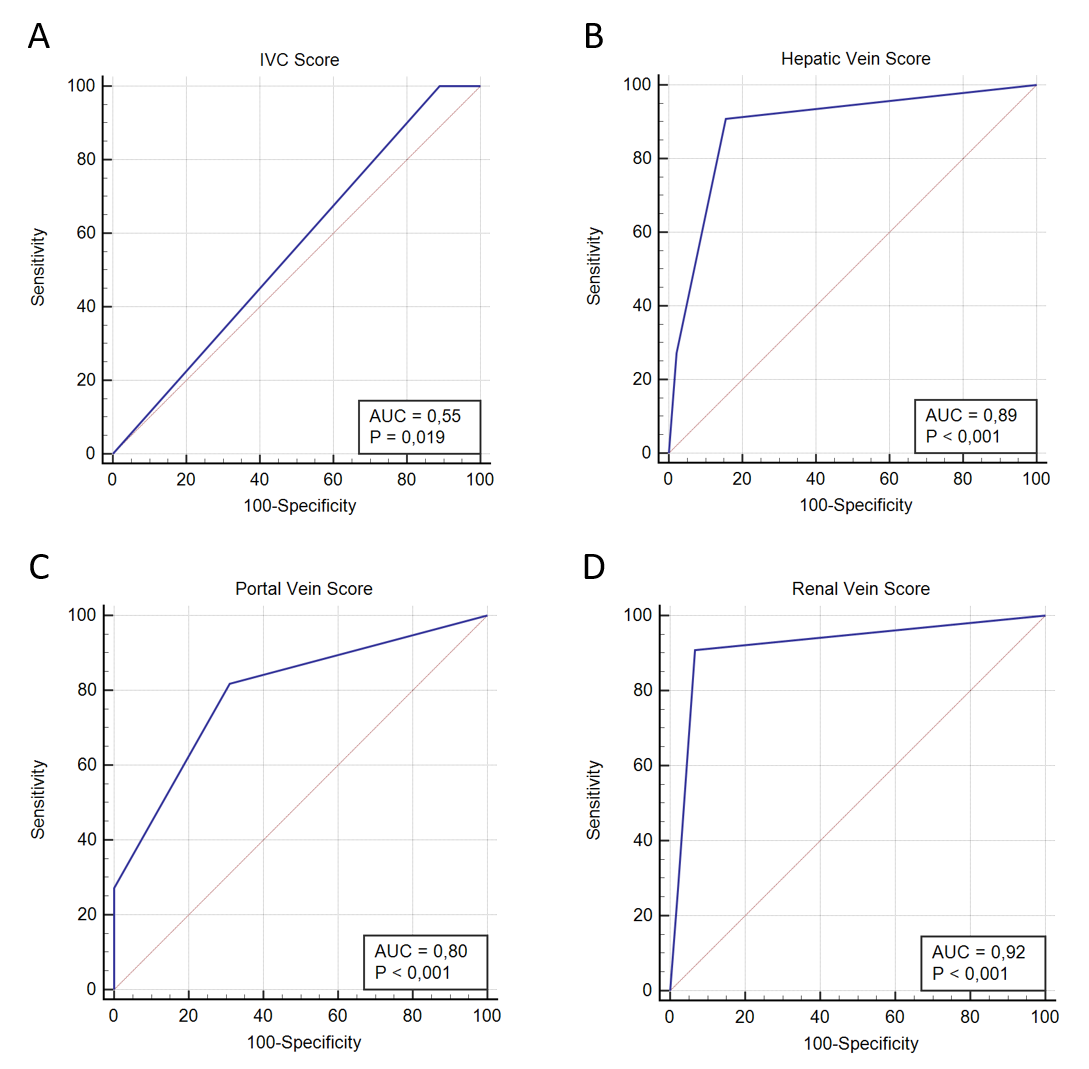

### **Figure S2.** **Receiver operating characteristic (ROC) curves for individual venous Doppler components in predicting elevated central venous pressure (CVP > 12 mmHg). (A) Inferior vena cava (IVC) score, AUROC = 0.55 (95% CI 0.42–0.69, *p* = 0.019); (B) hepatic vein score, AUROC = 0.89 (95% CI 0.77–0.96, *p* < 0.001); (C) portal vein score, AUROC = 0.80 (95% CI 0.67–0.89, *p* < 0.001); and (D) renal vein score, AUROC = 0.92 (95% CI 0.82–0.98, *p* < 0.001). Among individual sites, renal vein Doppler demonstrated the highest discriminative ability, followed by hepatic and portal veins, whereas the IVC score showed poor accuracy.**

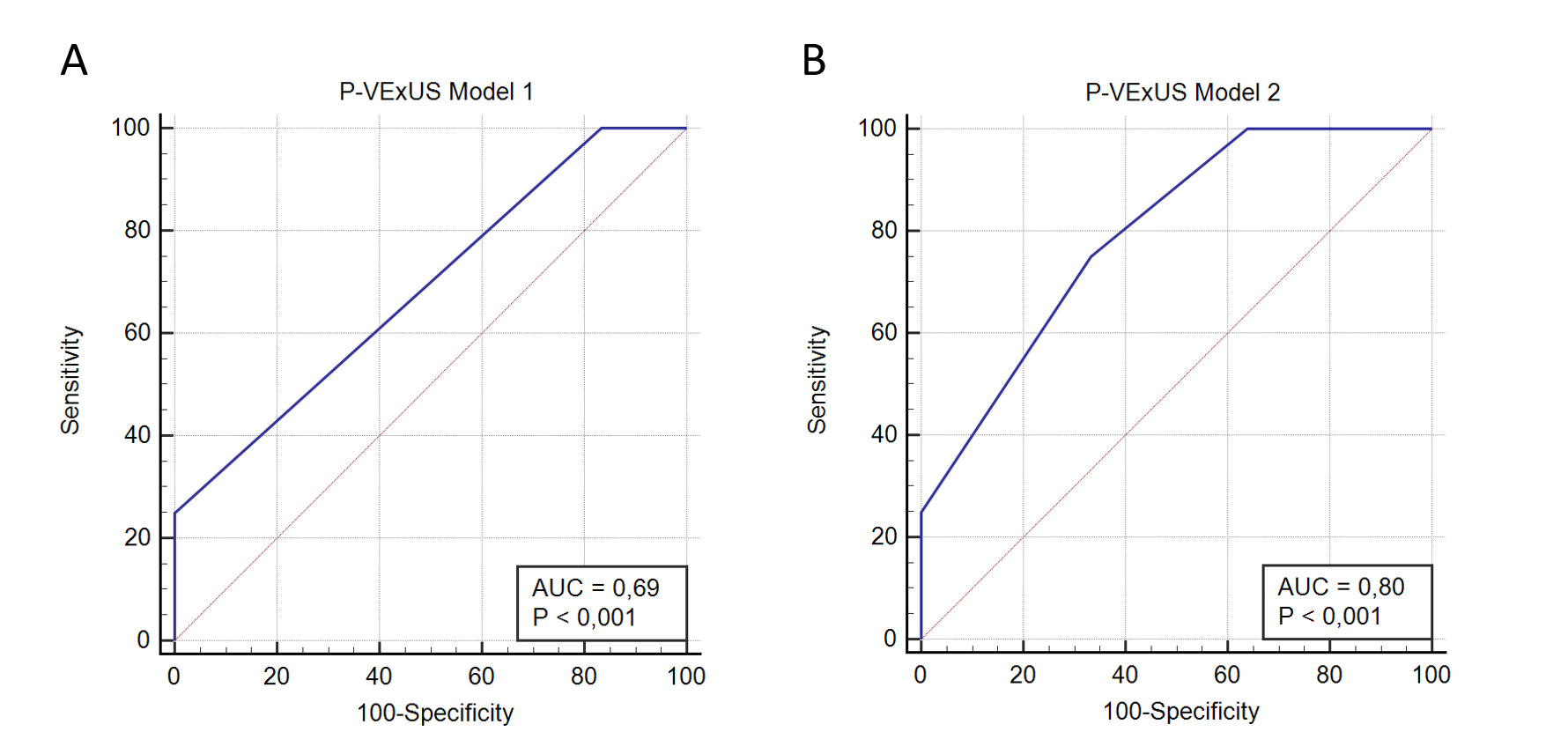

**Figure S3.** Receiver operating characteristic (ROC) curves for predicting low central venous pressure (CVP < 7 mmHg) using two pediatric Venous Excess Ultrasound Score (P-VExUS) models. (A) P-VExUS model 1 (categorical grading) demonstrated an AUROC of 0.69 (95% CI 0.55–0.81, *p* < 0.001). (B) P-VExUS model 2 (point-based score) achieved higher accuracy, with an AUROC of 0.80 (95% CI 0.67–0.89, *p* < 0.001). Model 2 showed significantly better discriminative performance compared to model 1 (*p* = 0.02).

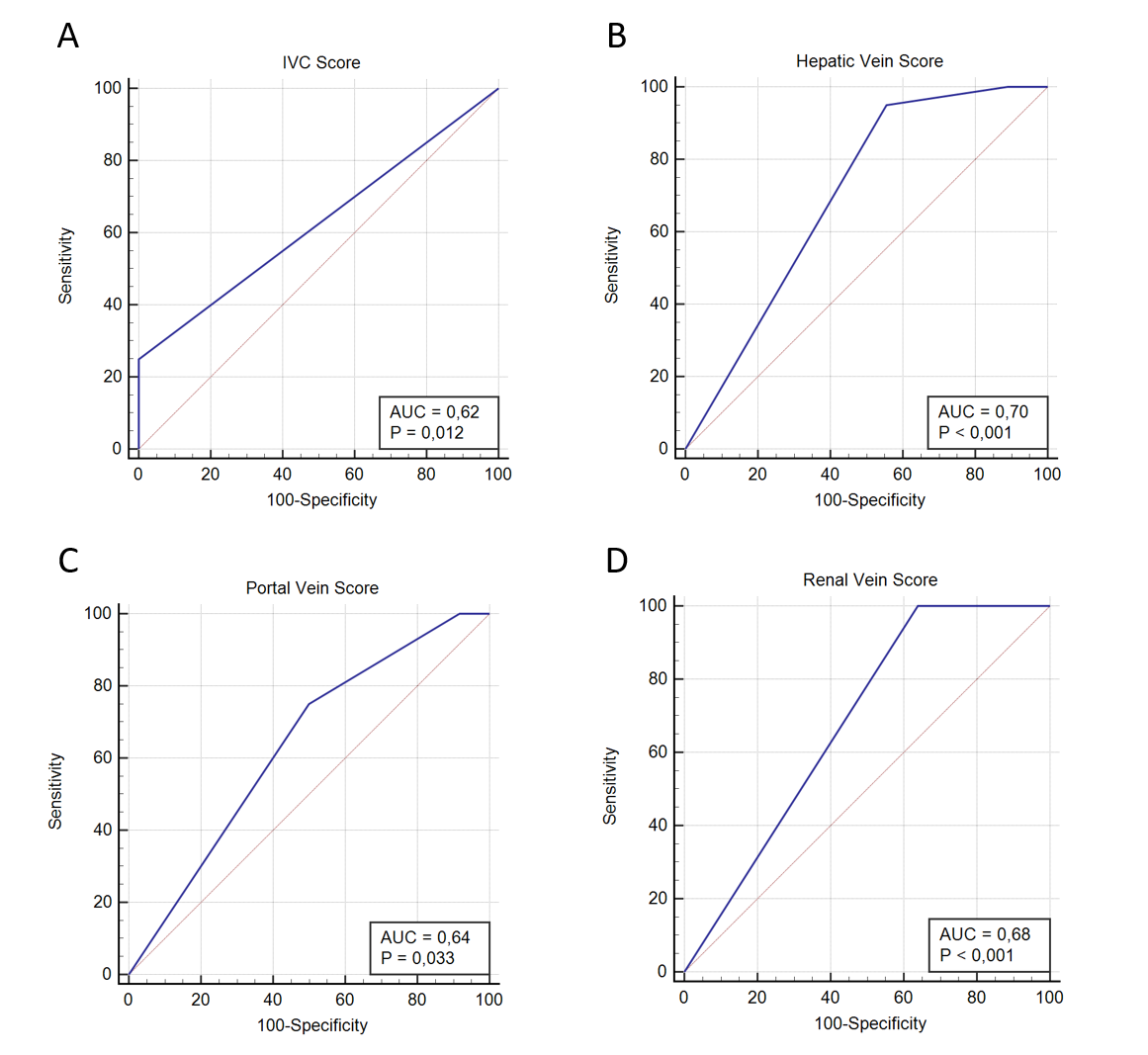

### **Figure S4. Receiver operating characteristic (ROC) curves of individual venous Doppler components for predicting low central venous pressure (CVP < 7 mmHg).** (A) Inferior vena cava (IVC) score, AUROC = 0.62 (95% CI 0.48–0.75, p = 0.012). (B) Hepatic vein score, AUROC = 0.70 (95% CI 0.56–0.81, p < 0.001). (C) Portal vein score, AUROC = 0.64 (95% CI 0.50–0.76, p = 0.033). (D) Renal vein score, AUROC = 0.68 (95% CI 0.54–0.80, p < 0.001).
